# ReCOVer study: A Cross-sectional Observational Study to Identify the *Re*habilitation Need in Post-discharge *COV*ID-19 Survivors

**DOI:** 10.1101/2021.04.19.21255750

**Authors:** Raktim Swarnakar, S. L. Yadav, V Srikumar, Kapil Dev Soni, Richa Aggrawal, Anjan Trikha

## Abstract

**Introduction:** With the increasing number of Coronavirus disease-2019 (COVID-19) cases there is simultaneous increase in recovered cases too. There are many post-covid complications where need for rehabilitation intervention is very conspicuous especially pulmonary, neurological complications. Hence data are of utmost importance to find out those rehabilitation needs among post-covid survivors.

**Methods and analysis:** ReCOVer (*Re*habilitation Need in Post-discharge *COV*ID-19 Survivors), a cross-sectional observational study protocol has been planned to find out rehab-need by assessing International Classification of Functioning, Disability and Health (ICF) core data set, COVID-19 Yorkshire Rehab Screen (C19-YRS) tool, The Post-COVID-19 Functional Status (PCFS) scale, barriers to functional independence and rehab services (affordability & availability). Post-discharge (minimum 1 weeks) Covid patients (required hospitalisation) will be included in the study. Study will be conducted through Telerehabilitation facility. Study will conform to the Strengthening the Reporting of Observational Studies in Epidemiology (STROBE) guidelines.

**Ethics and dissemination:** Study received ethical approval from Institute Ethics Committee, All India Institute of Medical Sciences (AIIMS), New Delhi, India. Findings will be disseminated at scientific conferences/meetings, peer-reviewed journals, and to relevant stakeholders including the ministry of health (if required).

## INTRODUCTION

Worldwide coronavirus disease (COVID-19) has affected 14 million and more than 8 million people have recovered from this disease.^1^ In India, confirmed cases crossed 11 lakhs and recovered cases crossed 7 lakhs.^2^ In the view of multiple post-COVID complications especially neurological and cardiopulmonary etc, rehabilitation need is very conspicuous. But currently evidence from literature and studies are lacking on rehabilitation of COVID-19 population. There is no study addressing rehab need in different subgroups of COVID-19 especially among different clinical severity of COVID-19 after discharge.

At this current scenario we propose to assess the need of rehabilitation in different clinical severity of COVID-19 patients after discharge from hospitalisation. This would form a basis for rehab recommendations for these patients and also it would help to address the knowledge gaps and to gather observations for future research.

## REVIEW OF LITERATURE

COVID-19, started in Wuhan, China in December, 2019, is caused by Severe acute respiratory syndrome coronavirus 2 (SARS-CoV-2). Then it spread to rest of the world causing a public health emergency. SARS-CoV-2 is a positive sense and single stranded RNA virus. Bat is the probable reservoir.

### Incubation period

2-14 days.

### Mode of transmission

Respiratory droplets.

### Clinical features

Fever, dry cough, fatigue, shortness of breath, sore throat are the common symptoms.

### COVID-19 confirmed case definition (WHO)^3^

A person with laboratory confirmation of COVID-19 infection, irrespective of clinical signs and symptoms.

### Clinical severity

According to WHO, it is classified as mild, moderate (pneumonia), severe (severe pneumonia) and critical stages (acute respiratory distress syndrome, sepsis, septic shock).

### Pathophysiology

SARS-CoV-2 enters human body via ACE2 receptor and primary this virus causes diffuse alveolar damage. Excessive immune reaction to the virus, causing cytokine storm, is mainly responsible for the clinical severity of COVID-19.

### Investigation

Confirmation is generally done by Real-time polymerase chain reaction in samples collected from upper and lower respiratory tract. Lymphopenia is a cardinal feature. Xray shows bilateral infiltrates, CT scan shows ground glass appearance.

### Treatment

^**4**^

### Mild

Antipyretics

### Moderate to severe

Supplemental oxygen therapy immediately to patients with SARI and respiratory distress, hypoxaemia or shock and target SpO2 > 94%. Empiric antimicrobials to treat all likely pathogens causing SARI and sepsis as soon as possible, within 1 hour of initial assessment for patients with sepsis.

### ARDS

Recognize severe hypoxemic respiratory failure when a patient with respiratory distress is failing to respond to standard oxygen therapy and prepare to provide advanced oxygen/ventilatory support. Implement mechanical ventilation using lower tidal volumes (4– 8 mL/kg predicted body weight, PBW) and lower inspiratory pressures (plateau pressure < 30 cmH2O). In adult patients with severe ARDS, prone ventilation for 12–16 hours per day is recommended.

### Septic shock

Recognize septic shock in adults when infection is suspected or confirmed AND vasopressors are needed to maintain mean arterial pressure (MAP) ≥ 65 mmHg AND lactate is ≥ 2 mmol/L, in absence of hypovolemia. Recognize septic shock in children with any hypotension (systolic blood pressure [SBP] < 5th centile or > 2 SD below normal for age) or two or more of the following: altered mental state; bradycardia or tachycardia (HR < 90 bpm or > 160 bpm in infants and HR < 70 bpm or > 150 bpm in children); prolonged capillary refill (> 2 sec) or feeble pulses; tachypnoea; mottled or cold skin or petechial or purpuric rash; increased lactate; oliguria; hyperthermia or hypothermia.

### Post-COVID-19 Complications

The most common clinical features were fever (99%), fatigue (70%), dry cough (59%), anorexia (40%), myalgias (40%), dyspnea (31%), and sputum production (27%).^5^

**Extra-respiratory manifestations of patients with COVID-19**^**6**^

- **Cardiac:** Acute cardiac injury (8–12%), heart failure (23–52%), arrhythmia (8.9– 16.7%), shock, acute myocarditis, chest tightness.
- **Gastrointestinal:** Anorexia (26.8%), diarrhoea (12.5%), nausea/vomiting (10.2%), abdominal pain/discomfort (9.2%).
- **Hepatic:** Abnormal aspartate aminotransferase or alanine aminotransferase values (16.1–53.1%).
- **Kidney:** Acute kidney injury (overall 0.5%; 2.9–23% in severe cases).
- **Neurological:** Dizziness (16.8%), headache (13.1%), skeletal muscle injury (10.7%), impaired consciousness (7.5%), acute cerebrovascular disease (2.8%), ataxia (0.5%), seizures (0.5%), meningoencephalitis, Guillain–Barré syndrome.
- **Olfactory and gustatory:** Hyposmia (5.1–20.4%), anosmia (79.6%), dysgeusia (8.5%), ageusia (1.7%).
- **Ocular:** Acute conjunctivitis (31.6%).
- **Cutaneous:** Erythematous rash (15.9%), hives rash (3.4%), vesicles (1.1%), acro-ischaemia, transient unilateral livedo reticularis.
- **Haematological:** Lymphopenia (56.5%), thrombocytopenia (16.4–32.3%), coagulation disorders, thrombotic events, antiphospholipid antibody.

It has been suggested that viral invasion of the central nervous system by SARS-CoV2 is possible by the synapse-connected route observed with other coronaviruses such as SARS-CoV and can lead to several neurological complications including ataxia, seizures, neuralgia, unconsciousness, acute cerebrovascular disease and encephalopathy.^7, 8^

The most common peripheral manifestation is hyposmia and Hyposmia (5.1–20.4%), anosmia (79.6%), dysgeusia (8.5%), ageusia (1.7%).^9^

Neuro-muscular complication as listed below:^10^

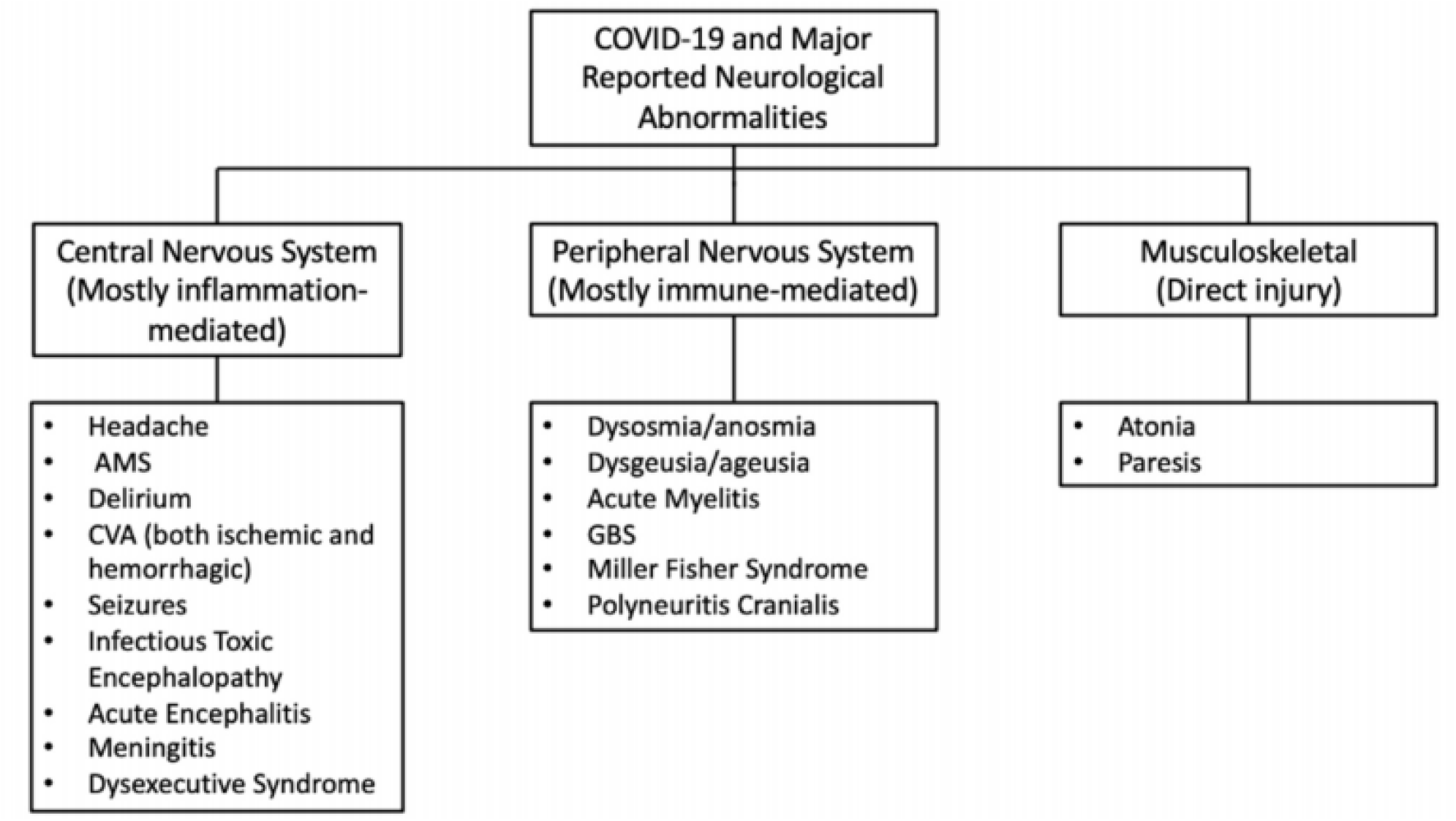

Helms et al.^11^ reported neurological complications in an observational case series of 58 patients admitted to the ICU for ARDS, secondary to COVID-19 in Strasbourg, France, between 3 March 2020 and 3 April 2020. Neurological findings were seen in 14% of patients at admission and 69% of cases were seen when they were weaned off sedation and paralytics. Most frequently observed symptoms were confusion (65%), agitation (69%), upper motor neuron syndrome signs like hyperreflexia with clonus and positive Babinski’s sign (69%) during the ICU stay, and a dysexecutive syndrome (33%) after discharge. MRI of the brain in patients who developed unexplained encephalopathic features revealed leptomeningeal enhancement (62%), perfusion abnormalities on MRI (100%), and ischemic CVA (23%). Only one out of the eight patients who underwent electroencephalogram (EEG) showed findings consistent with encephalopathy.

87.4% recovered COVID-19 patients reported persistence of at least 1 symptom, particularly fatigue (53.1%), dyspnea (43.4%), joint pain, (27.3%) and chest pain (21.7%).^12^

### Rehabilitation (current available evidences on COVID-19)

In India till now no studies have focussed rehabilitation in Covid. Currently rehabilitation is focussing on pulmonary rehabilitation in COVID-19 during hospital or ICU stay. Recently one consensus has been developed on post-Covid rehabilitation in UK,^13^ but in different settings it could be variable. For the time being, rehab guidelines for COPD has been recommended for rehabilitation of Covid patients. Most of the studies has emphasized pulmonary component but are lacking a comprehensive rehabilitation program. There is one study is going regarding telerehabilitation in Covid patient, but studies on rehab or even tele rehab in post-Covid population are lacking. One study^14^ has addressed 32 post-acute Covid patient with early rehabilitation in Italy.

### Outcome measure

#### C19-YRS tool^15^

It is recently developed, free but not validated. It is made for rehab need assessment in post Covid-19 patients. It addresses all domain of WHO ICF.

#### PCFS scale^16^

A recently developed scale, it covers the full spectrum of functional outcomes, and focuses on both limitations in usual duties/activities and changes in lifestyle in six scale grades. It is free to use, not validated.

#### ICF core data set^17^

For COVID-19 there is no separate ICF data set. Hence, in our study we would include all data in accordance with ICF rehabilitation data set including subcategories of musculoskeletal, neurological, cardiopulmonary condition according to individual patient’s condition as mentioned in ICF data set.

## RATIONALE OF STUDY

Worldwide more than 8 lakhs population have been recovered from Covid-19. Post-Covid neurological, cardiovascular sequelae have been identified, moreover it affects multiple system in our body. In this context rehabilitation would be an essential component in those patients for comprehensive management.

Hence, we propose this study to identify rehabilitation needs in these population for gathering information for further studies like type of rehabilitation intervention needed, to whom it would be needed, how can it be offered, its efficacy, and feasibility etc.

## AIM

To assess the rehabilitation-need in post-discharge COVID-19 survivors.

## OBJECTIVES

To determine the rehabilitation-need in post-discharge COVID-19 survivors by assessing:

### Primary objective

ICF core data set.

### Secondary objectives

1. COVID-19 Yorkshire Rehab Screen (C19-YRS) tool.
2. The Post-COVID-19 Functional Status (PCFS) scale.
3. Barriers to functional independence and rehab services (affordability & availability).

## MATERIALS & METHODS

- **Study setting:** The study will be conducted in the department of Physical Medicine and Rehabilitation (PM&R), AIIMS, New Delhi.
- **Study duration:** The study shall commence after approval from the institutional review board and the ethical committee and will be continued for a period of one month.
- **Type of study:** A cross-sectional observational study. Study will conform to the Strengthening the Reporting of Observational Studies in Epidemiology (STROBE) guidelines.
- **Study subjects:** Post-discharge COVID-19 positive patients.
- **Selection of cases:** As this is a sub-study of *“Post discharge outcomes of COVID-19 patients following admission to Intensive care unit: A prospective cohort study”* (**Ref. No IEC-291/17.04.2020, RP-11/2020**. Code No. A-COVID-41). The selection of the participants will be from the cohort of above mentioned study.
- **Follow-up:** No follow-up.
- **Sample size:** During COVID-19 pandemic, there is no study addressing rehabilitation need among post-discharge COVID-19 population. However, using 7 lakh recovered Covid-19 population in India till date, 5% margin of error, 95% confidence interval and 50% response distribution, sample size calculated was 384. We calculated our descriptive study sample size using CDC Epi Info™ 7.2.4.0 (StatCalc) software.
- **Inclusion criteria:**
  1. Age: any age group.
  2. Either gender.
  3. Post-discharge (minimum 1 week) of any confirmed COVID-19 cases those who required hospitalisation (required oxygenation, ICU monitoring or management, etc. for COVID-19).
- **Exclusion criteria:**
  1. No access to or unavailability of minimum phone-calling facility.
  2. Not willing to participate in the study.
- **Methodology:**
  1. **Patient selection:** From database according to the inclusion criteria.
  2. **Recruitment and Contacting the participants:** by telemedicine (telephonic calls).
  3. **Explanation and reassurance:** Patients will be explained about the whole procedure of the study.
  4. **Consent:** Consent will be taken according to the Telemedicine Practice Guidelines, India (2020).^18^
  5. **Participant’s details and history:** All demographic and clinical characteristics (age, sex, residence, source of contact of COVID-19 or contact tracing, duration of symptoms, durations of hospital admission, duration of ICU stay if any, history of diabetes, hypertension, kidney disease, cardiac disease or any other disease etc.), laboratory findings and medication history all will be collected from the ongoing cohort study (***Ref. No IEC-291/17*.*04*.*2020, RP-11/2020***. *Code No*. ***<u>A-COVID-41</u>***).
  6. **Addressing current clinical complaints (if any)**.
  7. **Data collection and assessment of C19-YRS tool and PCFS scale**: It will be done at single time-point (during telemedicine consultation).
  8. **Listing of barriers to functional independence, rehab services, hygiene practices:** For each barrier, its mechanism, coping strategies will be documented. Based on these policy implications would be addressed.
  9. **Information/education need:** Source of information or awareness regarding COVID-19, and in Yes/No format, knowledge regarding social distancing, hand hygiene, mask use will be documented.
  10. **Addressing support need:** Time given per day, income, and source of income of caregiver will be documented.
  11. **Listing of Physical activity participants doing (if any):** Types, intensity, duration of exercises/physical activity done per day.
- **Interventions:** Not applicable.

## Data Availability

This is a study protocol.

## STATISTICAL ANALYSIS

- Data will be entered in Microsoft EXCEL spreadsheet.
- Data will presented as mean ±SD/median (range, min, max) and frequency percentage.
- Stata 14.1 will be used in analysis.
- P value < 0.05 will be taken as statistically significant.

## Appendix

### APPENDIX-I Covid 19 Yorkshire Rehab Screen (C19-YRS)

Patient name and NHS number:

Time and date of call:

Staff member making call:

*We are getting in touch with people who have been discharged after having had a diagnosis of coronavirus disease (Covid-19). The purpose of this call is to find out if you are experiencing problems related to your recent illness with coronavirus. We will document this in your clinical notes. We will use this information to direct you to services you may need and inform the development of these services in the future*.

*This call will take around 15 minutes. If there’s any topics you don’t want to talk about you can stop the conversation at any point. Do you agree to talk to me about this today?* **Yes** □ **No** □

#### Opening questions

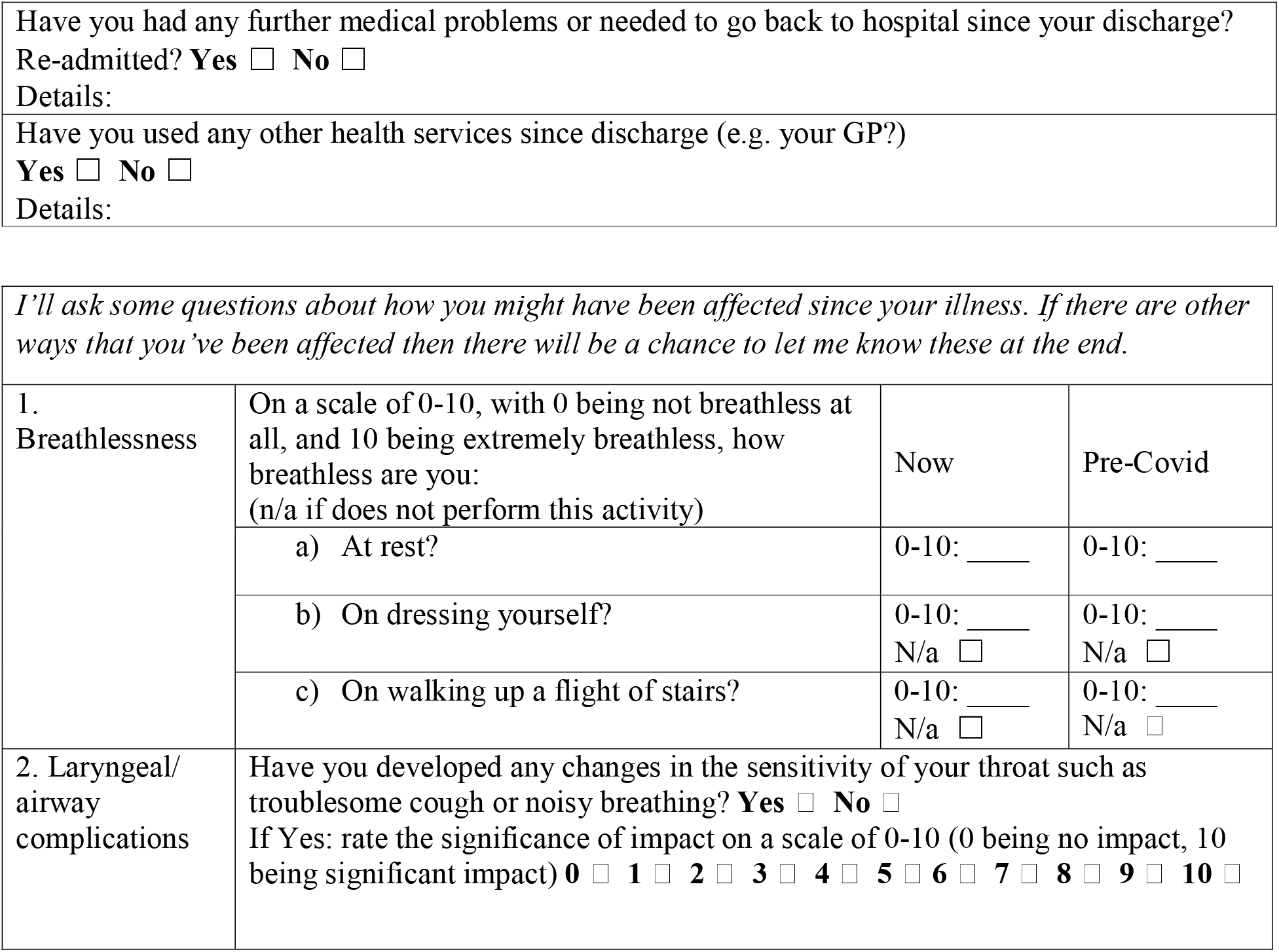

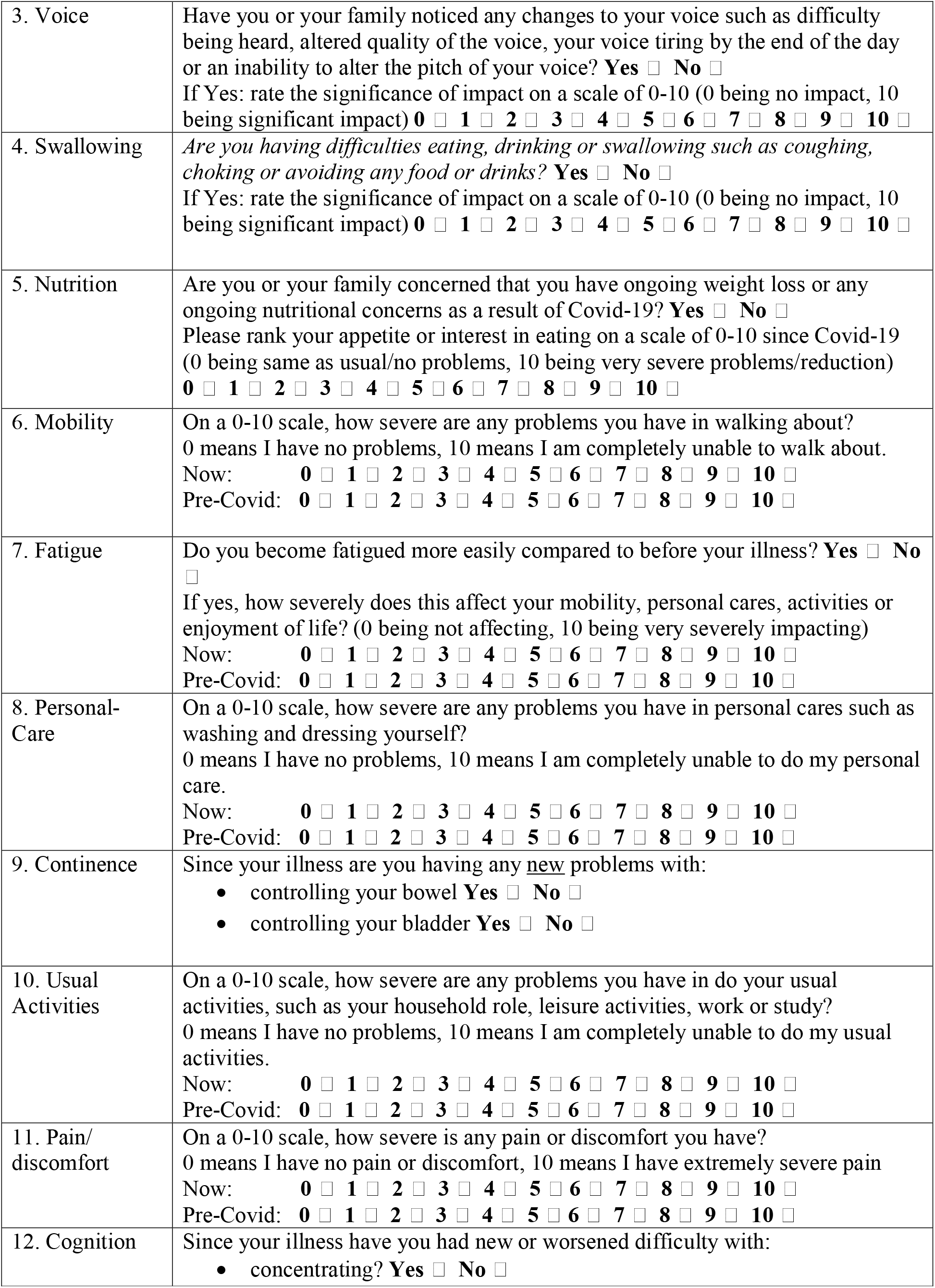

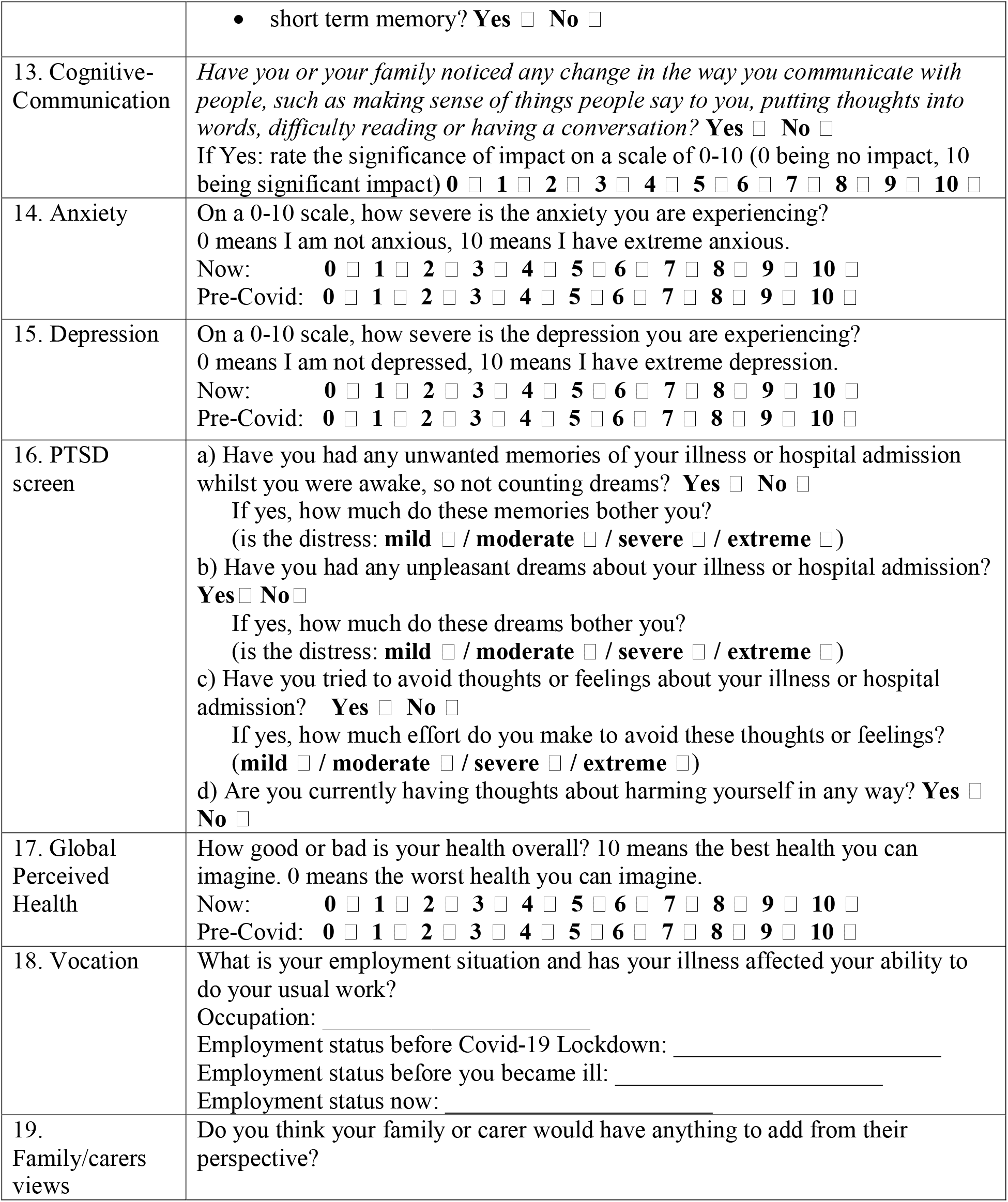

#### Closing questions

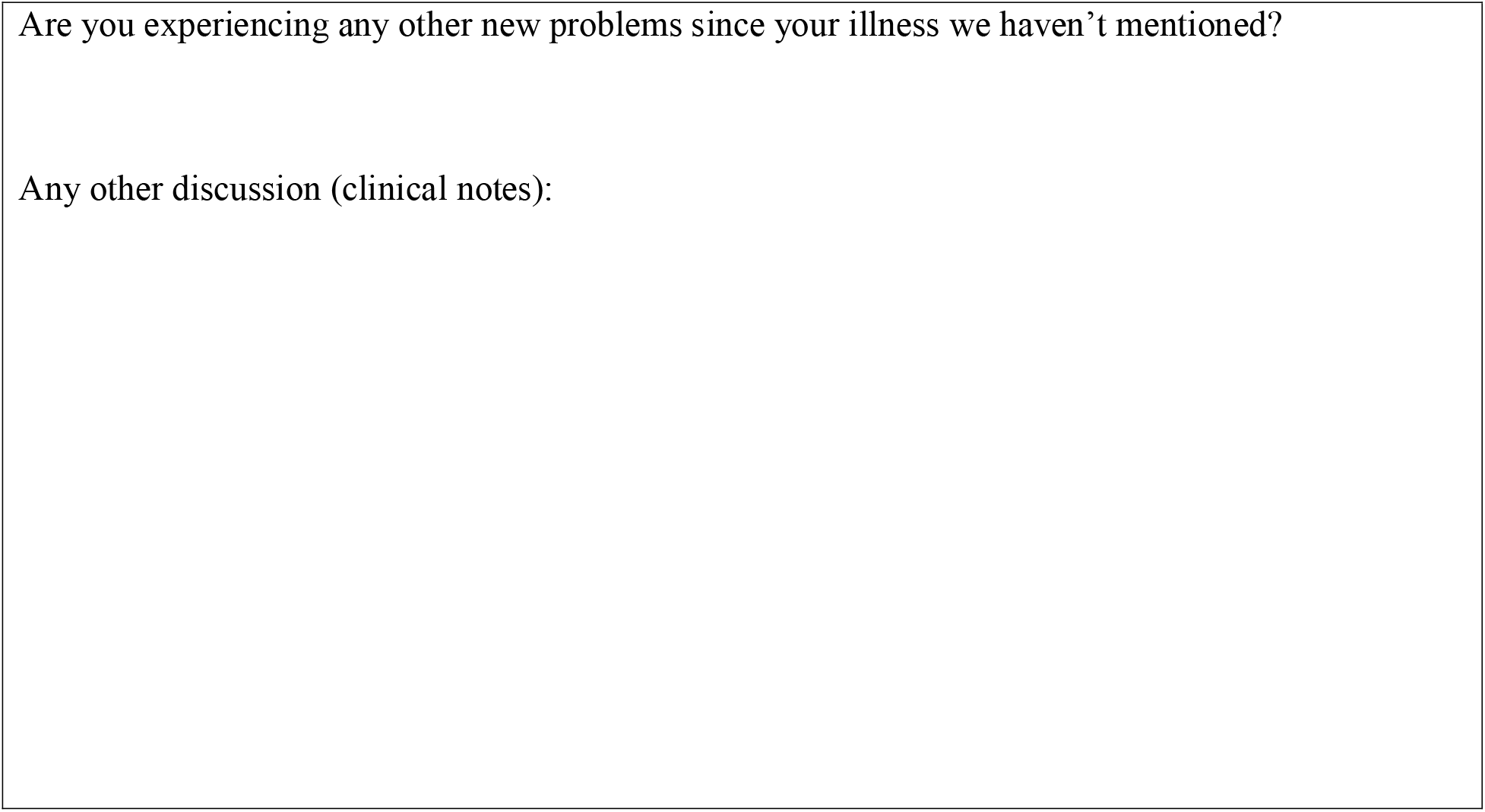

### APPENDIX-II Post-Covid Functional Status Scale

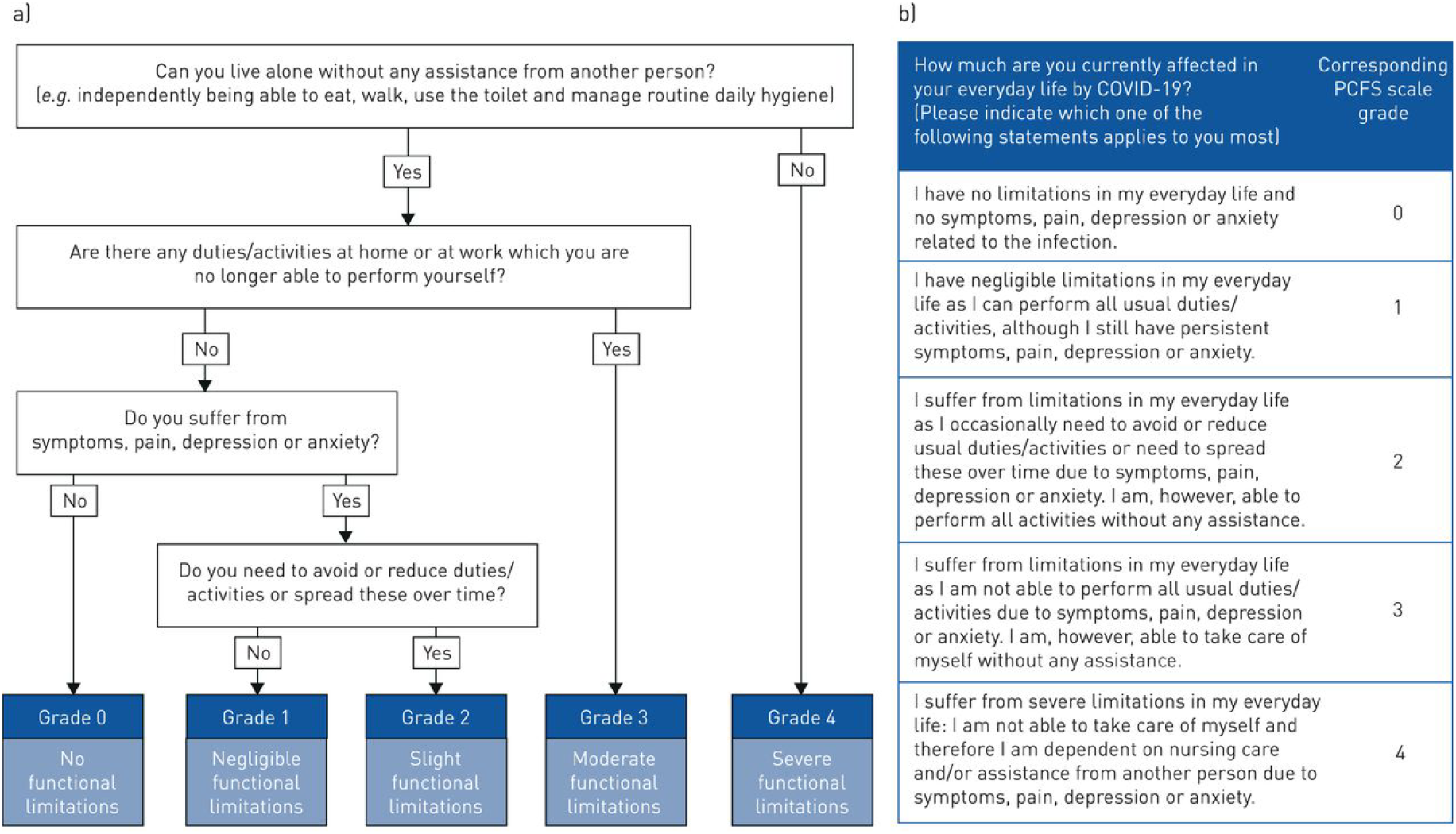

### APPENDIX-III ICF CORE DATA SET

https://www.icf-research-branch.org/icf-core-sets

